# Mapping the Network of Persistent Somatic Symptoms Across Diseases: A Longitudinal Analysis from the SOMACROSS Research Unit

**DOI:** 10.1101/2025.09.03.25334991

**Authors:** André Strahl, Kerstin Maehder, Anne Toussaint, Nele Hasenbank, Christoph Schramm, Ansgar W. Lohse, Meike Shedden-Mora, Tobias B. Huber, Stefan W. Schneider, Sonja Ständer, Yvonne Nestoriuc, Olaf von dem Knesebeck, Eik Vettorazzi, Antonia Zapf, Bernd Löwe

## Abstract

**Background:** Persistent Somatic Symptoms (PSS) are common, functionally disruptive, multifactorial, and often remain stable over time. Understanding how symptoms interact may clarify transdiagnostic patterns and inform treatment. This study investigated symptom networks in a transdiagnostic sample to identify central symptoms and assess temporal stability.

**Methods:** Patients (n=1134; 63.7% female; mean age 50.6±16.3 years) from the transdiagnostic SOMACROSS research unit were analysed with the Patient Health Questionnaire-15 (PHQ-15), modified Pain Disability Index (PDI), and two global items on overall symptom severity and impairment due to symptoms (EURONET-SOMA) at baseline and 6-month follow-up. Networks were estimated with regularised partial correlations (EBICglasso). Network stability and change were tested with bootstrap procedures and the Network Comparison Test (NCT).

**Results:** At baseline, four symptom clusters were identified: gastrointestinal, musculoskeletal pain, cardio-autonomic, and fatigue-sleep. Fatigue/low energy was a central symptom linking different network domains. Global symptom severity and impairment (EURONET-SOMA items) showed the strongest connectivity, reflecting their broad influence. Functional disability in home responsibilities and recreation (PDI items) acted as bridging nodes, connecting somatic symptoms with daily functioning. The follow-up network closely resembled the baseline structure. NCT confirmed temporal stability (global strength *p*=.815, structure invariance *p*=.180). Thus, both symptom clusters and central nodes remained consistent over six months.

**Conclusion:** Fatigue, overall symptom severity, impairment, and functional disability emerged as central drivers within network. Their stable centrality highlights them as possible intervention targets, suggesting that reducing fatigue and disability may interrupt self-reinforcing symptom cycles. These findings support a transdiagnostic view on PSS as a stable, interconnected system.

**Highlights:**

- Symptom clusters: gastrointestinal, musculoskeletal pain, cardio-autonomic and fatigue-sleep
- Fatigue and functional impairment were central symptom nodes across time points
- Global impairment and global symptom severity ratings were connected to somatic symptoms
- PSS network remained stable over 6 months, with no changes in strength or configuration

## Background

Persistent somatic symptoms (PSS) — physical complaints lasting several months or longer; regardless of their cause — are common, often disrupt daily functioning, and can substantially reduce quality of life, creating a considerable burden on healthcare systems [1, 2]. Most patients experience not just a single symptom, but clusters of symptoms, with physical and additionally psychological symptoms often occurring alongside [3, 4]. Once established, PSS tend to remain moderately stable for up to a decade [5], leading to impaired functioning, and an elevated utilisation of healthcare services [6, 7]. The underlying causes of PSS are multifactorial and comprise general biological, psychological and social factors in addition to disease-specific mechanisms [1]. Though prevalent, patients’ need for effective treatment is often unmet, as conventional biomedical care rarely addresses this complexity [2]. The clinical burden emphasises the importance of recognising PSS as a significant health challenge and developing improved strategies for early detection and intervention. Consequently, a transdiagnostic, multidisciplinary perspective has been advocated [1, 8, 9].

The SOMACROSS research unit (Persistent SOMAtic symptoms ACROSS diseases: From risk factors to modification, RU 5211) adopts this transdiagnostic approach, examining PSS within seven research projects across multiple diseases and patient cohorts [2]. The SOMACROSS RU quantifies PSS by integrating symptom severity and functional impairment using three well-established instruments: the Patient Health Questionnaire-15 (PHQ-15) [10], a modified version of the Pain Disability Index (PDI) [11, 12], and two global items developed within the EURONET-SOMA group assessing general symptom severity and impairment due to symptoms [13]. This approach reflects the biopsychosocial understanding of PSS [1] by capturing biological symptom expression as well as the psychosocial and functional impact.

Despite the progress that has been made in recognising the importance of PSS, several significant gaps in knowledge remain. One of these gaps concerns the latent structure of PSS, which refers to the underlying patterns and interrelationships between symptoms. This structure is clinically relevant as it could lead to more precise diagnostics and targeted treatments. Previous attempts to classify or group PSS using traditional factor analytic analyses or latent profile analysis have produced inconclusive results, primarily due to a lack of a stable and reproducible factor structure across patient populations [14-19]. Network analysis offers a promising alternative to examine and understand the complex interconnectedness between variables [20]. Senger et al [21] explored associations between different somatic symptoms, assessed with the 53 item Screening of Somatoform Disorder (SOMS-7T) and identified five main symptom cluster (neurological, gastrointestinal, urogenital, cardiovascular, musculoskeletal) in two independent inpatient and outpatient patients with somatoform disorders from a multicenter study [22]. However, network stability was limited by small sample size. While certain symptom groups appeared to demonstrate robust correlation, the nodes’ centrality, a measure of a symptom’s importance within the network, varied between the investigated samples.

Building on this method, the present study applies a network approach to examine the network structure of physical symptoms across medical conditions. It further investigates the temporal stability of PSS over two measurement points. This is particularly important as most questionnaires do not cover a period of several months and therefore persistence of symptoms cannot genuinely be verified. To our knowledge, no study has investigated the network structure of PSS over time. Accordingly, the aims of this study are (a) to investigate the structure of PSS, as defined by the SOMACROSS RU 5211, in a transdiagnostic sample of patients and (b) to observe the temporal development of symptoms using network analyses. This approach focuses on the complex interplay between PSS and subjective perception and functional impairment due to physical symptoms, aligning with the overarching goal of the SOMACROSS research framework, to identify symptoms that are centrally connected across various conditions, thereby contributing to the understanding of transdiagnostic symptom patterns [2].

## Methods

### Study design

This analysis used data from the interdisciplinary SOMACROSS RU [2]. Patients from a transdiagnostic cohort completed questionnaires on PSS and associated psychological constructs at baseline, 6 and 12 months (see study protocol [2]). The cohort comprised individuals with primary biliary cholangitis (PBC), primary sclerosing cholangitis (PSC), chronic kidney disease (CKD), ulcerative colitis (UC), irritable bowel syndrome (IBS), Chronic pruritus affecting primary non-lesional skin (CPNL), acute (ADa) and chronic atopic dermatitis (ADc), as well as somatic symptom disorder (SSD). Details are published elsewhere [23-27]. All studies received ethical approval, and written informed consent was obtained. For the present analysis, only patients with available 6-month follow-up data were included. Participants were excluded if >50% of PHQ-15 or PDI items, or either of the EURONET-SOMA items, were missing at baseline or follow-up.

### Outcome measures

PSS were operationalised with three stablished instruments. The PHQ-15 evaluates somatic symptom severity over the past four weeks, with 15 items graded on a 3-point Likert scale ranging from 0 (no impairment) to 2 (severe impairment), with a sum score (0–30 points) reflecting the subjective symptom severity (0–4 none/minimal; 5–9 low; 10–14 moderate; 15–30 high). Internal consistency is good (Cronbach’s α =.80 [10]. The item “menstrual cramps or other problems with your periods” was omitted to avoid gender-based bias. The adapted PDI measures the extent to which symptoms impair a person’s ability to participate in essential daily activities. The PDI, which typically evaluates the impact of pain [11], was adapted to focus on the broader impact of symptoms on daily life [12]. This instrument consists of seven items, scored on an 11-point scale ranging from 0 (‘no disability’) to 10 (‘worst disability’). Reliability and validity are well established [28, 29], with a Cronbach’s α ranging between.83 and.90 for the German version [30]. Additionally, the overall symptom severity was assessed using an 11-point scale (EURONET-SOMA1 item) ranging from 0 (‘no symptoms at all’) to 10 (‘worst possible symptoms’) over the past 7 days. Complementary, the EURONET-SOMA2 item assesses the overall impairment of patient’s daily life due to symptoms over the past week from 0 (‘no interference’) to 10 (‘complete interference’) [13]. **Table 1** shows the item abbreviations and content in the network.

**Table 1:** Item abbreviation and content.

| Abbreviation | Content |
| --- | --- |
| PHQ1 | Stomach pain |
| PHQ2 | Back pain |
| PHQ3 | Pain in arms, legs, or joints (knees, hips, etc.) |
| PHQ5 | Pain or problems during sexual intercourse |
| PHQ6 | Headaches |
| PHQ7 | Chest pain |
| PHQ8 | Dizziness |
| PHQ9 | Fainting spells |
| PHQ10 | Feeling your heart pound or race |
| PHQ11 | Shortness of breath |
| PHQ12 | Constipation, loose bowels, or diarrhea |
| PHQ13 | Nausea, gas, or indigestion |
| PHQ14 | Feeling tired or having low energy |
| PHQ15 | Trouble sleeping |
| PDI1 | Disability in family/home responsibilities |
| PDI2 | Disability in recreation |
| PDI3 | Disability in social activity |
| PDI4 | Disability in occupation |
| PDI5 | Disability in sexual behavior |
| PDI6 | Disability in self care |
| PDI7 | Disability in life-support activities |
| EURO1 | Global somatic symptom severity (EURONET-SOMA1) |
| EURO2 | Global somatic symptom interference (EURONET-SOMA2) |

### Statistical analysis

Continuous variables are expressed as mean (M) with standard deviation (SD), categorical variables as number and percentage. T-tests for dependent variables examined changes between the measurement points. Effect sizes were reported according to Cohen’s conventions: small (d =.20 –.49), medium (d =.50 –.79), and large (d ≥.80) effect [31]. Associations between variables were examined using Pearson correlations. Missing data was imputed using multiple imputation with predictive mean matching to retain full sample size for analysis [32]. Network computations were performed in R (version 4.5.1) and JASP (version 0.19.3), whereas descriptive statistics were calculated using IBM SPSS (version 20). Statistical significance was set at α =.05 (two-tailed).

#### Network analysis

Networks were estimated with R packages *bootnet* [33], *qgraph* [34], and *NetworkComparisonTest* [35]. All items were z-standardised (M = 0, SD = 1) to ensure comparability between the questionnaires. Regularised partial correlation networks (23 items: PHQ-15, PDI, EURONET-SOMA1/2) were estimated for each time point. Symptom are represented as a node, with edges between nodes indicating partial correlations. To obtain a robust, sparse network structure, Extended Bayesian Information Criterion Graphical LASSO (EBICglasso) algorithm was applied [36-38]. Centrality indices (*strength, closeness, betweenness, expected influence*) were computed [39]. *Strength* indicates how strongly a symptom is directly connected to others. *Closeness* reflects how fast a symptom can influence or be influenced by other symptoms and *betweenness* reflects how often a node serves as a bridge between two others. *Expected influence* considers the overall impact of an item to the network. Stability of edge weights was tested using bootstrapped intervals to assess sensitivity to sampling error. Finally, stability of network properties was examined via case-dropping bootstrap analysis (1000 iterations). According to guidelines, Centrality Stability (CS)-coefficients >.50 are considered sufficient for stable interpretation [33]. The Network Comparison Test (NCT) for dependent samples was used to test for differences between the baseline und FU network [35]. Based on 1000 permutations, the NCT evaluates global strength invariance (differences in overall connectivity, i.e., the sum of edge weights), network structure invariance (differences in the overall configuration of edges), edge invariance (differences in individual edge weights), and centrality invariance (differences in node centrality indices). To control for multiple testing, p-values were adjusted using the Benjamini–Hochberg false discovery rate (FDR) procedure.

### Sample size rational

Although the sample size in this study was fixed by the recruitment frameworks of the individual SOMACROSS projects, it remains relevant to consider whether this number is sufficient for reliable network estimation. In general, larger samples yield more accurate network models, enhancing the stability of the network [33, 40]. Traditional heuristics suggest 5 to 10 participants per variable as a general rule of thumb for correlation-based network models [33]. However, a recent simulation study has shown that this heuristic may not be sufficient. For networks comprising 20 to 30 nodes, sample sizes of approximately 300 to 500 participants are generally required to achieve acceptable sensitivity and specificity values under moderate effect sizes [41]. A Bootstrap analysis demonstrated that correlation estimates stabilise with increasing sample size, with the critical point for a correlation of r =.1 at n = 470 [42]. In accordance with these recommendations, the number of patients in this study is considered sufficient for the proposed network analysis. Subgroup analyses by individual diseases were not performed due to limited sample sizes.

## Result

### Sample characteristics and longitudinal changes

Between September 2021 and February 2024, 1343 patients were recruited. Of these, 1208 provided follow-up questionnaires at 6 months. After exclusion of 74 patients with >50% missing responses, the final sample comprises 1134 patients. For these participants, 0.2% of remaining missing values were imputed. The final sample comprised 218 patients with PBC (19.2%), 204 patients with PSC (18.0%), 208 patients with CKD (18.3%), 115 patients with UC (10.1%), 105 patients with IBS (9.3%), 34 patients with CPNL (3.0%), 8 patients with ADa (0.8), 31 patients with ADc (2.7) and 211 patients with SSD (18.6%). Participants were predominantly female (63.7%; n = 722), with a mean age of 50.6 (SD 16.3) years and mean BMI of 26.1 (SD 5.8) kg/m^2^. Most reported no migration background (81.6%) and had completed twelve or more (59.5%) or ten years (26.4%) of formal school education. At baseline, the mean PHQ-15 score was 8.5 (SD 5.4), with 40.9% participants scoring values ≥10, indicating at least moderate symptom severity. Complete sociodemographic and clinical characteristics stratified by disease group are presented in **Table 2**.

**Table 2:** Sociodemographic characteristics and clinical baseline of participants.

| Variable | Overall<br>(n = 1134) | PBC<br>(n = 218) | PSC<br>(n = 204) | CKD<br>(n = 208) | UC<br>(n = 115) | IBS<br>(n = 105) | ADa/ADc<br>(n = 39) | CPNL<br>(n = 34) | SSD<br>(n = 211) |
| --- | --- | --- | --- | --- | --- | --- | --- | --- | --- |
| Age in years [mean (SD)] | 50.6 (16.3) | 60.1 (10.6) | 45.1 (13.1) | 61.1 (14.8) | 39.9 (12.4) | 40.9 (15.5) | 47.0 (22.4) | 67.0 (11.6) | 44.6 (13.8) |
| Gender (n, %) |  |  |  |  |  |  |  |  |  |
| Male | 412, 36.3% | 17, 7.8% | 126, 61.8% | 118, 56.7% | 31, 27.0% | 23, 21.9% | 18, 46.2% | 11, 32.4% | 68, 32.2% |
| Female | 720, 63.5% | 201, 92.2% | 78, 38.2% | 90, 43.3% | 83, 72.2% | 81, 77.1% | 21, 53.8% | 23, 67.6% | 143, 67.8% |
| Diverse | 2, 0.2% | --- | --- | --- | 1, 0.9% | 1, 1.0% | --- | --- | --- |
| Body mass index [mean (SD)] | 26.1 (5.8) | 27.2 (5.4) | 25.1 (4.7) | 27.4 (5.8) | 25.0 (5.6) | 23.4 (4.3) | 24.5 (3.8) | 27.0 (5.5) | 27.0 (7.0) |
| Migration background<sup>‡</sup> (n, %) |  |  |  |  |  |  |  |  |  |
| Yes | 207, 18.3% | 43, 19.7% | 28, 13.7% | 51, 24.5% | 14, 12.2% | 20, 19.0% | 6, 15.4% | 4, 11.8% | 41, 19.4% |
| No | 925, 81.6% | 174, 79.7% | 176, 86.3% | 157, 75.0% | 101, 87.8% | 85, 81.0% | 32, 82.1% | 30, 88.2% | 170, 80.6% |
| missing | 2, 0.2% | 1, 0.5% | --- | --- | --- | --- | 1, 2.6% | --- | --- |
| Formal school education (n, %) |  |  |  |  |  |  |  |  |  |
| ≤ 9 years | 154, 13.6% | 52, 23.9% | 18, 8.8% | 45, 21.6% | 3, 2.6% | --- | 4, 10.3% | 10, 29.4% | 22, 10.4% |
| 10 years | 299, 26.4% | 75, 34.4% | 59, 28.9% | 48, 23.1% | 21, 18.3% | 29, 27.6% | 8, 20.5% | 16, 47.1% | 43, 20.4% |
| 12+ years | 675, 59.5% | 91, 41.7% | 126, 61.8% | 111, 53.4% | 91, 79.1% | 76, 72.4% | 26, 66.7% | 8, 23.5% | 146, 69.2% |
| Other | 5, 0.4% | --- | 1, 0.5% | 4, 1.9% | --- | --- | --- | --- | --- |
| missing | 1, 0.1% | --- | --- | --- | --- | --- | 1, 2.6% | --- | --- |
| PHQ-15 score [mean (SD)] | 8.5 (5.4) | 8.0 (5.4) | 5.3 (4.1) | 6.6 (5.0) | 11.0 (4.2) | 11.3 (3.8) | 5.2 (3.2) | 6.8 (4.5) | 12.1 (5.1) |
| PHQ-15 symptom severity level |  |  |  |  |  |  |  |  |  |
| none/minimal (0-4) | 303, 26.7% | 67, 30.7% | 103, 50.5% | 85, 40.9% | 6, 5.2% | 1, 1.0% | 19, 48.7% | 12, 35.3% | 10, 4.7% |
| low (5-9) | 366, 32.3% | 74, 33.9% | 64, 31.4% | 69, 33.2% | 37, 32.2% | 35, 33.3% | 16, 41.0% | 11, 32.4% | 60, 28.4% |
| moderate (10-14) | 304, 26.8% | 46, 21.1% | 32, 15.7% | 35, 16.8% | 50, 43.5% | 50, 47.6% | 3, 7.7% | 9, 26.5% | 79, 37.4% |
| high (15-30) | 161, 14.2% | 31, 14.2% | 5, 2.5% | 19, 9.1% | 22, 19.1% | 19, 18.1% | 1, 2.6% | 2, 5.9% | 62, 29.4% |
| Modified PDI score [mean (SD)] | 22.8 (15.9) | 18.5 (15.3) | 13.6 (13.5) | 17.0 (14.0) | 30.6 (12.2) | 31.0 (9.6) | 19.8 (16.4) | 21.5 (16.8) | 34.3 (14.1) |
| EURONET-SOMA-1 [mean (SD)] | 4.3 (2.7) | 3.8 (2.4) | 2.8 (2.3) | 3.2 (2.6) | 5.2 (2.1) | 6.0 (1.9) | 4.2 (2.7) | 4.9 (3.0) | 6.13 (2.0) |
| EURONET-SOMA-2 [mean (SD)] | 4.1 (2.9) | 3.5 (2.6) | 2.5 (2.5) | 2.9 (2.7) | 5.0 (2.2) | 5.9 (1.9) | 4.0 (3.0) | 4.6 (2.9) | 6.15 (2.3) |

<sup>‡</sup> defined as individuals where they themselves or at least one parent was not born in Germany; Ada: acute atopic dermatitis; ADc: chronic atopic dermatitis; CKD: chronic kidney disease; CPNL: Chronic pruritus affecting primary non-lesional skin; IBS: irritable bowel syndrome; modified PDI: modified Pain Disability Index; PBC: primary biliary sclerosis; PHQ-15: Patient Health Questionnaire-15; PSC: primary sclerosing cholangitis; SD: Standard deviation; SSD: somatic symptom disorder, UC: ulcerative colitis

Over six months, PSS scores showed small but significant improvements. The mean PHQ-15 value decreased by 0.5 points ([95%CI 0.3-0.7]; T_(1133)_ = 4.420, *p* <.001, d =.13). The PDI declined from 22.8 (SD 15.9) at baseline to 21.8 (SD 15.8) at follow-up (ΔM 1.0 [95% CI 0.4-1.6]; T_(1133)_ = 3.181, *p* =.002, d =.09). The perceived overall symptom severity (EURONET-SOMA1) decreased from 4.3 (SD 2.6) to 4.1 (2.6; ΔM 0.2 [95% CI 0.06-0.32]; T_(1133)_ = 2.885, *p* =.004, d =.09). Similarly, the overall impairment due to symptoms (EURONET-SOMA2) declined from 4.1 (SD 2.8) to 3.9 (SD 2.9; ΔM 0.2 [95% CI 0.06-0.34]; T_(1133)_ = 2.741, *p* =.006, d =.08).

At baseline, strong correlations were observed between both EURONET-SOMA items, while the PDI was moderately associated with both and EURONET-SOMA measures. The PHQ-15 showed only moderate correlations with the other three scales. All correlations were statistically significant (*p* <.001). Similar correlation patterns were found at 6 months-follow-up (**Table 3**), suggesting a stable relationship between the components of PSS over time. These stable patterns of associations between PSS components provided the basis for subsequent item-level network analyses.

**Table 3:**
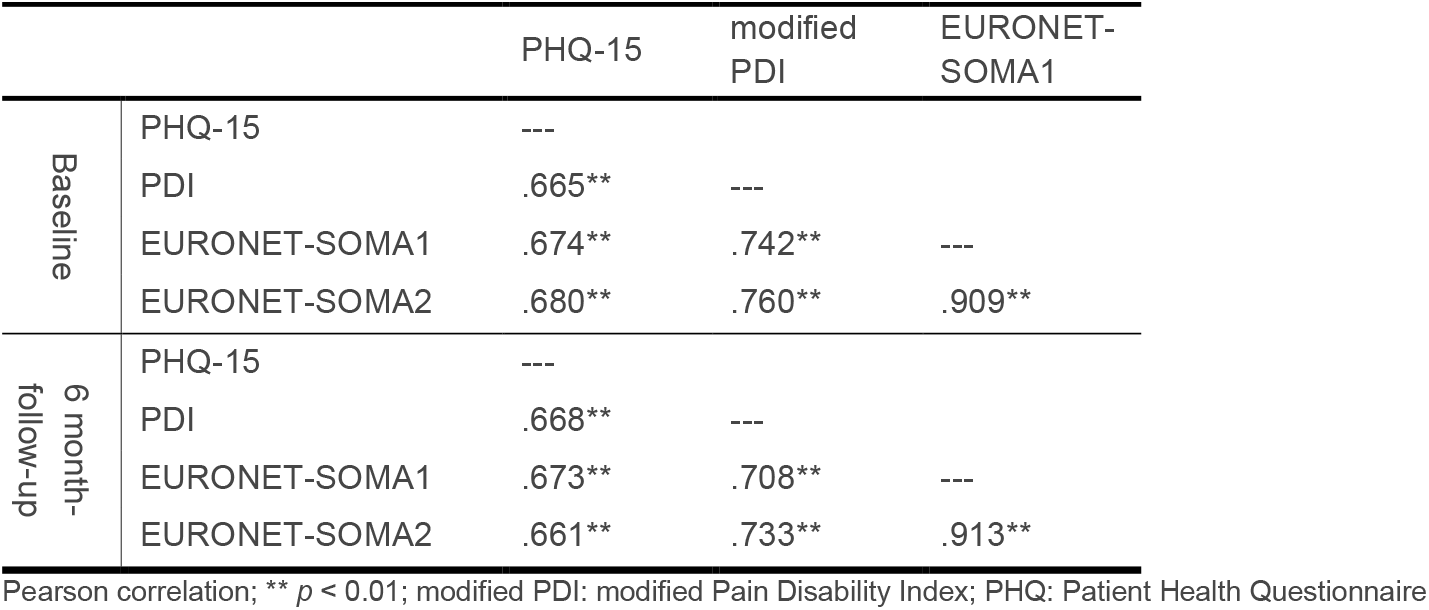
Bivariate correlations between components of persistent somatic symptoms at baseline and follow-up (n = 1134)

|  |  | PHQ-15 | modified PDI | EURONET-SOMA1 |
| --- | --- | --- | --- | --- |
| Baseline | PHQ-15 | --- |  |  |
|  | PDI | .665** | --- |  |
|  | EURONET-SOMA1 | .674** | .742** | --- |
|  | EURONET-SOMA2 | .680** | .760** | .909** |
| 6 month-<br>follow-up | PHQ-15 | --- |  |  |
|  | PDI | .668** | --- |  |
|  | EURONET-SOMA1 | .673** | .708** | --- |
|  | EURONET-SOMA2 | .661** | .733** | .913** |

Pearson correlation; \*\* $p < 0.01$ ; modified PDI: modified Pain Disability Index; PHQ: Patient Health Questionnaire

### Transdiagnostic network analysis to examine the interconnectedness of persistent somatic symptoms

#### Baseline network structure

At baseline, the combined symptom network (23 nodes) comprised a moderately sparse structure with 130 non-zero edges out of 253 possible (density = 51.4%; sparsity =.486, mean edge weight =.038; **Figure 1A**). Several coherent symptoms clusters were evident. Gastrointestinal symptoms (stomach pain [PHQ1], nausea [PHQ13], bowel irregularities [PHQ12]) and musculoskeletal pain (back pain [PHQ2], extremity/joint pain [PHQ3]) formed distinct clusters. A cardio-autonomic cluster (chest pain [PHQ7], palpitations [PHQ10], shortness of breath [PHQ11], dizziness [PHQ8], reflected the co-occurrence of cardiopulmonary complaints. In addition, fatigue/low energy [PHQ14] and sleep disturbance [PHQ15] formed another prominent cluster. Within the domain of functional impairment, family/home responsibilities [PDI1], recreation [PDI2], and social activity [PDI3] were highly interconnected, with disability in occupation [PDI4] contributing to a broad functional impairment cluster. The two global EURONET-SOMA items demonstrated the strongest interconnection of all nodes.

**Figure 1:**
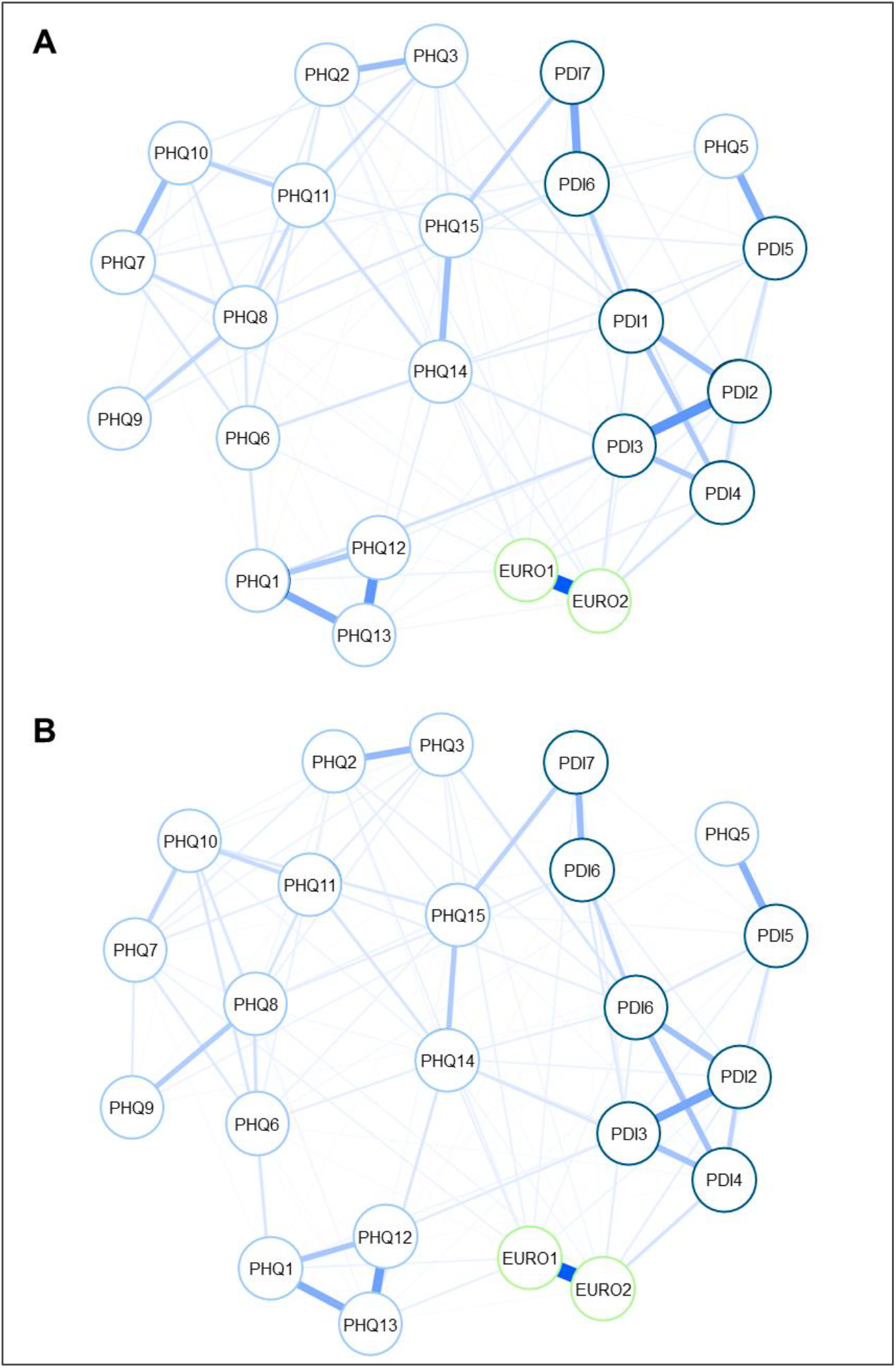
Estimated network plot for somatic symptoms and impairment at baseline and 6-month follow-up. Estimated network plot for persistent somatic symptoms in the total sample. **(A)** Baseline network graph and **(B)** 6-month follow-up network graph, each with regularized partial correlations among z-standardized symptom and disability items at baseline (N = 1134 patients). The network includes 14 Patient Health Questionnaire-15 (PHQ-15) symptom items (excluding the menstrual item), seven adapted Pain Disability Index items (PDI1–PDI7), and two global somatic impact items from the EURONET-SOMA project (EURO1, EURO2). Edges represent partial correlation coefficients estimated using the EBICglasso (Extended Bayesian Information Criterion graphical LASSO) method; thicker edges denote stronger associations.

#### Baseline centrality indices

Centrality analyses indicate the significance of specific nodes within the network (**Figure 2**). The EURONET-SOMA items emerged as the most central nodes, showing the highest *strength* (EURO1: z =1.32; EURO2: z = 1.73) and *expected influence* (EURO1: z = 1.45; EURO2: z = 1.86). Despite their broad connectivity, *betweenness* and *closeness* were comparatively low, suggesting they integrate symptoms globally without serving as short-path bridges. The network structure also prominently featured the PDI items. Both, disabilities in family/home responsibilities [PDI1] and recreation [PDI2] resulted as key networks hubs. In particular, PDI1 demonstrated high *betweenness* (z = 2.43), *closeness* (z = 1.87), as well as above-average *strength* (z = 1.05). Therefore, this item represented an important bridge connecting functional disability to somatic symptom clusters. Within the PHQ symptoms, fatigue/low energy [PHQ14; *strength* z =.71, *expected influence* z =.84] emerged as particularly central, linking somatic complaints with functional disability. By contrast, dizziness, fainting spells, or sexual problems were rather peripheral. Notably, while stomach pain [PHQ1] showed only average *strength*, its high *betweenness* suggests that this node may act as a bridge, linking otherwise distinct symptom clusters despite not being equally severe in all participants.

**Figure 2:**
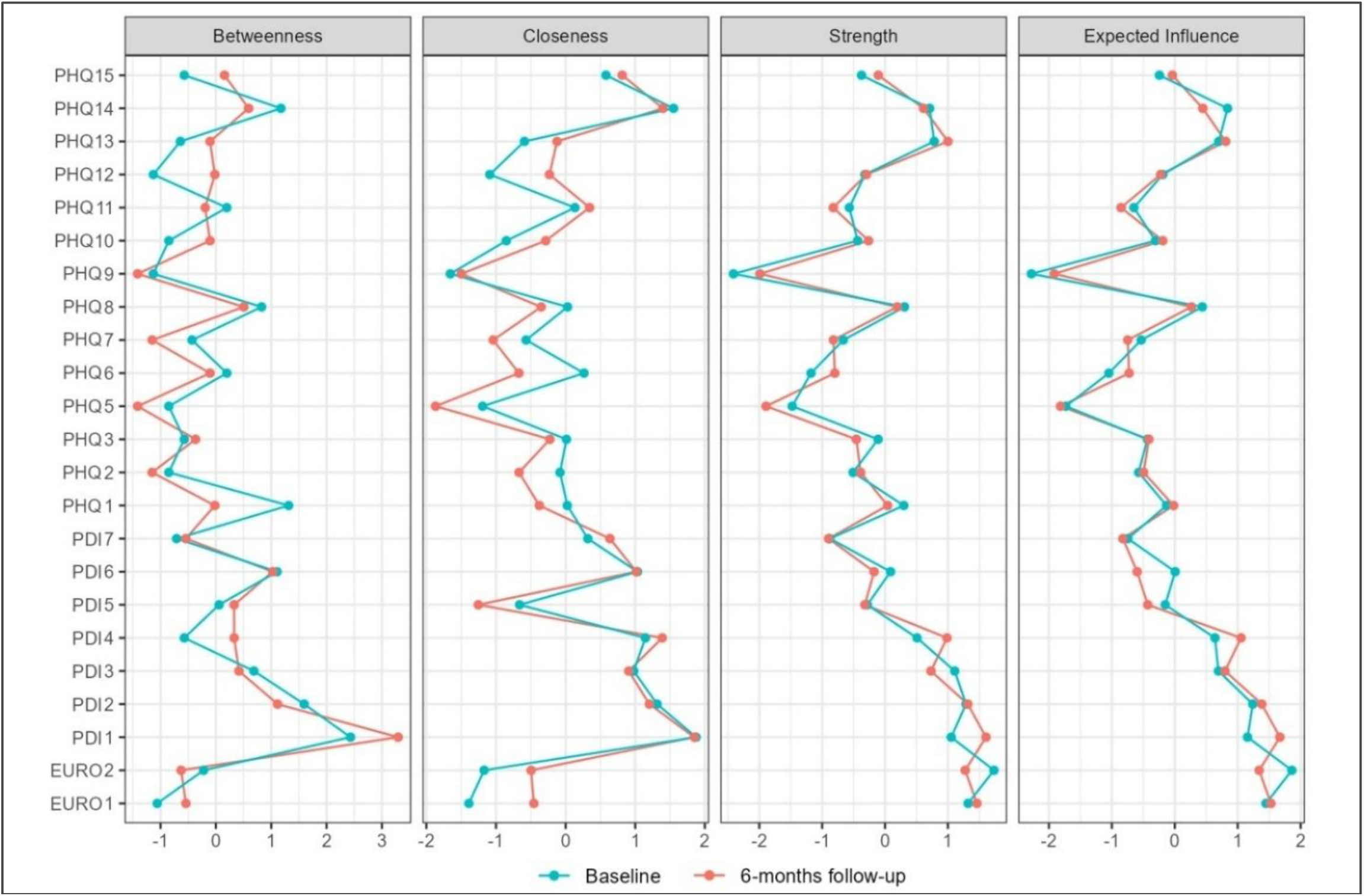
Centrality measures for network plots at baseline and 6-month follow-up. Comparison of the centrality measures (betweenness, closeness, strength, and expected influence) for the 23 items of the baseline and 6-month follow-up network. Displayed are standardized centrality metrics (z-scores) for each node in the symptom network. Description for PHQ1-PHQ15, PDI1-PDI7, EURO1-EURO2 see **Table 1**.

#### Network stability analysis at baseline

At baseline, the network showed overall robust stability of centrality indices (**Supplementary Fig. S1**). Case-dropping bootstrap analysis yielded a CS-coefficient of.75 for both *strength and expected influence*. These indices remain stable even after randomly removing up to 75% of participants. *Closeness* also showed acceptable stability (CS =.594), whereas the CS-coefficient for *betweenness* was markedly lower (CS = 0.439). Bootstrapped difference tests revealed that several of the strongest edges were significantly stronger than weaker ones, confirming the robustness of key connections in the network.

### Transdiagnostic symptom network over time

#### Network structure and centrality indices at 6-month follow-up

The follow-up network included 132 non-zero edges out of 253 (density = 52.2%; sparsity =.478, mean edge weight =.038; **Figure 1B**). The overall structure closely resembled the baseline network, with gastrointestinal [PHQ1, PHQ12, PHQ13], musculoskeletal [PHQ2, PHQ3], and cardio-autonomic symptoms (PHQ7, PHQ8, PHQ10, PHQ11] forming distinct symptom clusters. Fatigue/low energy [PHQ14] and sleep disturbance [PHQ15] interconnected and embedded within the broader network. Within the disability domain, PDI1 to PDI4 remained strongly interconnected, while the two EURONET-SOMA global items maintained their position as the strongest connected cluster. The centrality pattern closely mirrored the baseline results. Fatigue/low energy [PHQ14] persisted as one of the most central somatic symptoms, showing high *strength* (z = 0.61) and *closeness* (z = 1.4) and the two EURONET-SOMA items still ranked among most central nodes [EURO1; *strength* z = 1.46, *expected influence* z = 1.53; EURO2; *strength* z = 1.23, *expected influence* z = 1.34). Full centrality estimates are presented in **Figure 2**. Bootstrap case-dropping analysis confirmed high stability for *strength* and *expected influence* (CS =.750), while *closeness* showed acceptable stability (CS =.517). Also at follow-up, *betweenness* again demonstrated reduced robustness (CS =.205) (**Supplementary Figure S2**).

#### Network comparison

At 6-month follow-up, the network structure closely resembled the baseline network and showed a comparable degree of overall connectivity. All symptom clusters persisted across both time points. Minor shifts in node centrality indices were observed. A permutation-based NCT for dependent groups demonstrated no evidence for global or structural change of the symptom network from baseline to 6 months. Global *expected influence* was comparable across time (S =.026; *p* =.815). The network structure invariance test was non-significant (M =.110; *p* =.180). Edge-wise comparisons yielded no differences after FDR correction (all adjusted *p* ≥ 0.85). Centrality invariance tests also indicated no differences for *strength, expected influence, bridge strength*, or *bridge expected influence* (all *p* ≥ 0.88; range 0.878–1.000). These results indicate that the PSS network’s overall connectivity, topology, and node importance remained stable over 6 months.

## Discussion

This study applied a network analysis approach in a transdiagnostic cohort of 1134 patients to investigate symptom interactions at baseline and 6-month follow-up. The network emphasises three interrelated domains regarding PSS: (a) interactions between individual symptoms, (b) global appraisals of overall symptom severity and impairment, and (c) functional disability across daily life domains. The multimodal assessment considers not only the presence and severity of symptoms, but also their effect on daily life and functioning. Including both reflects the dimensional nature of PSS, in line with contemporary definitions that emphasize this multifaceted nature [1, 2]. The results suggest that global and functional impairments are not merely ‘downstream’ outcomes, but constituent elements of the network, interacting closely with individual somatic symptoms. Fatigue (loss of energy) emerged as one of the most central somatic complaint, showing strong connections with both other symptoms and disability. Likewise, global ratings of symptom severity and impairment (EURONET-SOMA) and functional disability (PDI) ranked among the most central elements, underscoring their pivotal role in linking symptoms with daily functioning in the PSS network.

The analysis identified four symptom clusters – gastrointestinal, musculoskeletal pain, cardio-autonomic symptoms and a fatigue-sleep cluster – also described in the recent literature [1, 21, 43]. While previous studies primarily identified these clusters through factor-analytic approaches, the present findings demonstrate that they also emerge in a network framework across diverse somatic conditions. This convergence suggests that these clusters are stable and transdiagnostically valid. Taken together, these results suggest that core symptom patterns reoccur across different methods, diagnoses and populations. Furthermore, the overall network structure of symptom–symptom relationships remained stable over 6 months, reflecting the persistent nature of PSS [1, 5].

### Symptom interconnectedness and centrality

The bivariate correlations indicate that somatic symptom severity, symptom-related disability and overall symptom severity and impairment are closely interrelated at baseline and after six months. However, the estimated networks at both time points demonstrated domain-specific clustering, consistent with known somatic symptom domains, and cross-domain connectivity mediated by key connector symptoms. In particular, fatigue acts as a common pathway across diverse conditions. These findings are consistent with a prior network study that identified tiredness/exhaustion as a central node [21]. The results also align with clinical experience emphasizing the presence of fatigue in various chronic illnesses [44].

Another important finding is the central role of overall symptom severity and impairment due to symptoms in the PSS network. The two EURONET-SOMA items were strongly connected to nearly all somatic symptoms. Rather than being downstream outcomes, they operated as constituent network nodes, as proposed [2]. Thus, increased global symptom burden and impairment are associated with a broader and more intense spectrum of individual symptoms, and vice versa. This resonates with prior evidence demonstrating significant associations between somatic symptom severity and lower quality of life, and functional impairment [45, 46]. This network analysis extends this knowledge by illustrating how global perceptions co-occur with specific symptom-to-symptom connections. The strong centrality of the EURONET-SOMA items illustrates their broad connectivity, while PDI items formed a cohesive cluster linked to symptom nodes, emphasizing the relationship between functional limitations and symptom experience.

The bootstrap analyses to evaluate network stability (CS-coefficient) indicate robust stability of *strength, closeness*, and *expected influence*, confirming the reliability of these indices. The lower stability of *betweenness* represents a potential limitation. However, this is a common finding in psychological networks, where *betweenness* tends to be less reliably [39].

### Temporal Stability of PSS

A key question in understanding PSS is whether symptom interrelations remain stable over time. This is particularly important because questionnaires refer to a specific period of time (e.g. the PHQ-15 to the last 4 weeks [10]), while the term “persistent” itself implies several months. Although dependent t-tests indicate statistically significant score changes over time, effect sizes were very small, ranging from.08 to.13. The network analyses confirmed the temporal stability of PSS, as the overall configuration of strong symptom-to-symptom connections remained consistent from baseline to 6-month follow-up. This aligns with research demonstrating the chronic course of PSS [5, 47, 48]. The results provide, for the first time, empirical evidence that symptoms not only remain present over months but also continue to be interconnected within the network. The consistent centrality of fatigue and functional impairment across both networks suggests that, once established, a network of persistent symptoms and disabilities can sustain itself in the absence of major interventions or external changes. This aligns with the theory that symptoms can form feedback loops that sustain the overall syndrome. In PSS, initial triggers can cause symptom networks (e.g., fatigue, pain, worry) that persist even after the trigger resolves [1].

### Clinical Implications

The shared symptom network structure despite varying diagnoses suggests that persistent symptoms interact comparably once established, supporting a unified approach to PSS classification and treatment focused on shared symptom patterns [49]. This finding offers empirical support for a generic PSS construct and highlight the central role of symptoms like fatigue, often considered secondary in disease-specific management. From a theoretical standpoint, this highlights the potential for PSS to be understood as an emergent network phenomenon, with similar patterns of symptom connectivity developing across individuals and diagnoses, probably driven by shared biopsychosocial mechanisms. Currently, management of PSS is challenged by diagnostic uncertainty and limited treatment strategies [50]. Identifying central nodes offers practical guidance for intervention. As an example, fatigue can result in decreased activity and social engagement, leading to greater disability. Consequently, reduced activity can worsen fatigue and amplify attention to bodily complaints, further contributing to symptoms such as pain or sleep problems. The conceptualization is congruent with cognitive-behavioral models of PSS, which describe vicious cycles wherein symptoms, distress, and disability reinforce each other [51]. Therefore, these results promote an integrated biopsychosocial, interdisciplinary care that includes medical, psychological, and physiotherapeutic support [9]. These implications align with proposed evidence-based treatment principles for PSS, which emphasize person-centred communication and biopsychosocial explanation, complemented by targeted psychological interventions (e.g., CBT, psychodynamic or mindfulness-based approaches), pharmacological support for comorbid disorders, and multimodal interdisciplinary care in severe cases [1].

### Strengths and Limitations

This study has clear strengths: It is the first to investigate the network of PSS in a prospective design, incorporating two measurement points, enabling analysis of temporal stability. Further strength includes the large sample size (n = 1134), the transdiagnostic design, and the use of established and standardised instruments. Including patients with different underlying medical conditions and functional syndromes, enhances the generalisability of the results. The application of regularized partial correlation network and bootstrap techniques ensure reliable network estimation [36]. However, limitations should be considered. Causality cannot be inferred. Therefore, central symptoms may not represent primary drivers, as associations may be bidirectional or confounded. While including functional impairment is a strength, the network’s mix of symptoms and disability measures adds complexity. Strong connections between somatic symptoms and disability may partly reflect overall severity rather than the specific causal paths. Furthermore, subgroup analyses by diagnosis were not feasible due to sample size constraint. Although the absence of subgroup analyses may have precluded the identification of disease-specific network alterations, the focus on a broad patient sample permits the characterization of common PSS components. Another limitation concerns the interpretation of temporal stability. While the NCT suggest that symptoms and their interconnections remain stable, this stability may partly reflect consistent reporting styles or trait-like illness perceptions rather than unchanged symptom experiences. Future studies including clinician ratings, ecological momentary assessment, or objective markers could help disentangle true symptom persistence.

## Conclusion

In conclusion, this study demonstrated that persistent somatic symptoms form a highly interconnected and temporally stable network across diverse medical conditions. Fatigue, overall symptom severity and impairment due to symptoms as well as disabilities in family/home responsibilities and recreation emerged as consistently central nodes. The persistence of network structure over six months provides empirical confirmation that PSS are enduring and remain stable interconnected over time. These findings support the conceptualization of PSS a dynamic, self-reinforcing system rather than isolated complaints. These insights contribute to a transdiagnostic understanding of PSS and highlight the relevance of symptom interconnectivity for diagnosis and treatment.

## Data Availability

All data produced in the present study are available upon reasonable request to the authors

## Acknowledgement

The authors would like to thank Prof. Dr. med. Gudrun Schneider for her valuable support during the initial phase of project acquisition and data collection. This work was carried out within the framework of Research Unit 5211 (FOR 5211) ‘Persistent SOMAtic Symptoms ACROSS Diseases: From Risk Factors to Modification (SOMACROSS)’, funded by the German Research Foundation (Deutsche Forschungsgemeinschaft, DFG). The DFG project number for the coordination project is 445297796 (speaker: Professor Bernd Löwe, MD); see also https://gepris.dfg.de/gepris/projekt/445297796.

## References

[1] B. Lowe, A. Toussaint, J.G.M. Rosmalen, W.L. Huang, C. Burton, A. Weigel, J.L. Levenson, P. Henningsen, Persistent physical symptoms: definition, genesis, and management, Lancet 403(10444) (2024) 2649–2662.

[2] B. Lowe, V. Andresen, O. Van den Bergh, T.B. Huber, O. von dem Knesebeck, A.W. Lohse, Y. Nestoriuc, G. Schneider, S.W. Schneider, C. Schramm, S. Stander, E. Vettorazzi, A. Zapf, M. Shedden-Mora, A. Toussaint, Persistent SOMAtic symptoms ACROSS diseases - from risk factors to modification: scientific framework and overarching protocol of the interdisciplinary SOMACROSS research unit (RU 5211), BMJ Open 12(1) (2022) e057596.

[3] S. Kohlmann, B. Gierk, A. Hilbert, E. Brahler, B. Lowe, The overlap of somatic, anxious and depressive syndromes: A population-based analysis, J Psychosom Res 90 (2016) 51–56.

[4] P. Husing, A. Smakowski, B. Lowe, M. Kleinstauber, A. Toussaint, M.C. Shedden-Mora, The framework for systematic reviews on psychological risk factors for persistent somatic symptoms and related syndromes and disorders (PSY-PSS), Front Psychiatry 14 (2023) 1142484.

[5] S. Atasoy, P. Henningsen, H. Sattel, J. Baumert, I.M. Ruckert-Eheberg, U. Kraus, A. Peters, K.H. Ladwig, C. Hausteiner-Wiehle, Stability and predictors of somatic symptoms in men and women over 10 years: A real-world perspective from the prospective MONICA/KORA study, J Psychosom Res 162 (2022) 111022.

[6] M.L. Joustra, K.A. Janssens, U. Bultmann, J.G. Rosmalen, Functional limitations in functional somatic syndromes and well-defined medical diseases. Results from the general population cohort LifeLines, J Psychosom Res 79(2) (2015) 94–9.

[7] L.M. McAndrew, L.A. Phillips, D.A. Helmer, K. Maestro, C.C. Engel, L.M. Greenberg, N. Anastasides, K.S. Quigley, High healthcare utilization near the onset of medically unexplained symptoms, J Psychosom Res 98 (2017) 98–105.

[8] B. Lowe, S. Zipfel, O. van den Bergh, P. Henningsen, Reconsidering Persistent Somatic Symptoms: A Transdiagnostic and Transsymptomatic Approach, Psychother Psychosom 94(1) (2025) 20–25.

[9] A. Toussaint, A. Weigel, B. Lowe, E.-S. group, The overlooked burden of persistent physical symptoms: a call for action in European healthcare, Lancet Reg Health Eur 48 (2025) 101140.

[10] K. Kroenke, R.L. Spitzer, J.B. Williams, The PHQ-15: validity of a new measure for evaluating the severity of somatic symptoms, Psychosom Med 64(2) (2002) 258–66.

[11] C.A. Pollard, Preliminary validity study of the pain disability index, Percept Mot Skills 59(3) (1984) 974.

[12] R. Mewes, W. Rief, N. Stenzel, H. Glaesmer, A. Martin, E. Brahler, What is “normal” disability? An investigation of disability in the general population, Pain 142(1-2) (2009) 36–41.

[13] W. Rief, C. Burton, L. Frostholm, P. Henningsen, M. Kleinstauber, W.J. Kop, B. Lowe, A. Martin, U. Malt, J. Rosmalen, A. Schroder, M. Shedden-Mora, A. Toussaint, C. van der Feltz-Cornelis, E.-S. Group, Core Outcome Domains for Clinical Trials on Somatic Symptom Disorder, Bodily Distress Disorder, and Functional Somatic Syndromes: European Network on Somatic Symptom Disorders Recommendations, Psychosom Med 79(9) (2017) 1008–1015.

[14] M. Witthoft, W. Hiller, N. Loch, F. Jasper, The latent structure of medically unexplained symptoms and its relation to functional somatic syndromes, Int J Behav Med 20(2) (2013) 172–83.

[15] C.H. Tsai, Factor analysis of the clustering of common somatic symptoms: a preliminary study, BMC Health Serv Res 10 (2010) 160.

[16] P. Fink, T. Toft, M.S. Hansen, E. Ornbol, F. Olesen, Symptoms and syndromes of bodily distress: an exploratory study of 978 internal medical, neurological, and primary care patients, Psychosom Med 69(1) (2007) 30–9.

[17] M. Eliasen, T. Jorgensen, A. Schroder, T.M. Dantoft, P. Fink, C.H. Poulsen, N.B. Johansen, L.F. Eplov, S. Skovbjerg, S. Kreiner, Somatic symptom profiles in the general population: a latent class analysis in a Danish population-based health survey, Clin Epidemiol 9 (2017) 421–433.

[18] M. Witthoft, S. Fischer, F. Jasper, F. Rist, U.M. Nater, Clarifying the latent structure and correlates of somatic symptom distress: A bifactor model approach, Psychol Assess 28(1) (2016) 109–15.

[19] P. Bos, R. Monden, C. Benraad, J. Groot, R. Oude Voshaar, D. Hanssen, Latent profile analysis of biopsychosocial measures in older patients with (un)explained persistent somatic symptoms, Compr Psychiatry 135 (2024) 152527.

[20] D. Borsboom, M.K. Deserno, M. Rhemtulla, S. Epskamp, E.I. Fried, R.J. McNally, D.J. Robinaugh, M. Perugini, J. Dalege, G. Costantini, A.-M. Isvoranu, A.C. Wysocki, C.D. van Borkulo, R. van Bork, L.J. Waldorp, Network analysis of multivariate data in psychological science, Nature Reviews Methods Primers 1(1) (2021).

[21] K. Senger, J. Heider, M. Kleinstauber, M. Sehlbrede, M. Witthoft, A. Schroder, Network Analysis of Persistent Somatic Symptoms in Two Clinical Patient Samples, Psychosom Med 84(1) (2022) 74–85.

[22] M. Kleinstauber, C. Allwang, J. Bailer, M. Berking, C. Brunahl, M. Erkic, H. Gitzen, M. Gollwitzer, J.M. Gottschalk, J. Heider, A. Hermann, C. Lahmann, B. Lowe, A. Martin, J. Rau, A. Schroder, J. Schwabe, J. Schwarz, R. Stark, F.D. Weiss, W. Rief, Cognitive Behaviour Therapy Complemented with Emotion Regulation Training for Patients with Persistent Physical Symptoms: A Randomised Clinical Trial, Psychother Psychosom 88(5) (2019) 287–299.

[23] B. Lowe, Y. Nestoriuc, V. Andresen, E. Vettorazzi, A. Zapf, S. Hubener, K. Maehder, L. Peters, A.W. Lohse, Persistence of gastrointestinal symptoms in irritable bowel syndrome and ulcerative colitis: study protocol for a three-arm randomised controlled trial (SOMA.GUT-RCT), BMJ Open 12(6) (2022) e059529.

[24] Y. Nestoriuc, F. Pauls, K. Blankenburg, S. Hahn, H. Wittenbecher, B. Lowe, A. Toussaint, Modifiable factors for somatic symptom persistence in patients with somatic symptom disorder: study protocol for a longitudinal cohort with an embedded ecologically momentary assessment (SOMA.SSD), BMJ Open 14(11) (2024) e083500.

[25] M.C. Shedden-Mora, B. Jessen, C. Schmidt-Lauber, B. Lowe, M. Rosch, H. Dannemeyer, J. Gloy, O. Van den Bergh, T.B. Huber, Predictors of somatic symptom persistence in patients with chronic kidney disease (SOMA.CK): study protocol for a mixed-methods cohort study, BMJ Open 12(11) (2022) e067821.

[26] A. Toussaint, L. Buck, J. Hartl, B. Lowe, C. Schramm, Factors associated with severity and persistence of fatigue in patients with primary biliary cholangitis: study protocol of a prospective cohort study with a mixed-methods approach (SOMA.LIV), BMJ Open 12(12) (2022) e061419.

[27] G. Schneider, S. Stander, S. Kahnert, M.P. Pereira, C. Mess, V. Huck, K. Agelopoulos, G. Frank, S.W. Schneider, Biological and psychosocial factors associated with the persistence of pruritus symptoms: protocol for a prospective, exploratory observational study in Germany (individual project of the Interdisciplinary SOMACROSS Research Unit [RU 5211]), BMJ Open 12(7) (2022) e060811.

[28] R.C. Tait, C.A. Pollard, R.B. Margolis, P.N. Duckro, S.J. Krause, The Pain Disability Index: psychometric and validity data, Arch Phys Med Rehabil 68(7) (1987) 438–41.

[29] R.C. Tait, J.T. Chibnall, S. Krause, The Pain Disability Index: psychometric properties, Pain 40(2) (1990) 171–182.

[30] U. Dillmann, P. Nilges, H. Saile, H.U. Gerbershagen, PDI: Pain Disability Index - deutsche Fassung, ZPID (Leibniz Institute for Psychology) – Open Test Archive, 2011.

[31] J. Cohen, A power primer, Psychol Bull 112(1) (1992) 155–9.

[32] R.J.A. Little, Missing-Data Adjustments in Large Surveys, Journal of Business & Economic Statistics 6(3) (1988).

[33] S. Epskamp, D. Borsboom, E.I. Fried, Estimating psychological networks and their accuracy: A tutorial paper, Behav Res Methods 50(1) (2018) 195–212.

[34] S. Epskamp, A.O.J. Cramer, L.J. Waldorp, V.D. Schmittmann, D. Borsboom, qgraph: Network Visualizations of Relationships in Psychometric Data, Journal of Statistical Software 48(4) (2012).

[35] C.D. van Borkulo, R. van Bork, L. Boschloo, J.J. Kossakowski, P. Tio, R.A. Schoevers, D. Borsboom, L.J. Waldorp, Comparing network structures on three aspects: A permutation test, Psychol Methods 28(6) (2023) 1273–1285.

[36] S. Epskamp, E.I. Fried, A tutorial on regularized partial correlation networks, Psychol Methods 23(4) (2018) 617–634.

[37] G.G.R. Leday, S. Richardson, Fast Bayesian inference in large Gaussian graphical models, Biometrics 75(4) (2019) 1288–1298.

[38] D.R. Williams, Bayesian Estimation for Gaussian Graphical Models: Structure Learning, Predictability, and Network Comparisons, Multivariate Behav Res 56(2) (2021) 336–352.

[39] L.F. Bringmann, T. Elmer, S. Epskamp, R.W. Krause, D. Schoch, M. Wichers, J.T.W. Wigman, E. Snippe, What do centrality measures measure in psychological networks?, J Abnorm Psychol 128(8) (2019) 892–903.

[40] G. Costantini, S. Epskamp, D. Borsboom, M. Perugini, R. Mõttus, L.J. Waldorp, A.O.J. Cramer, State of the aRt personality research: A tutorial on network analysis of personality data in R, Journal of Research in Personality 54 (2015) 13–29.

[41] M.A. Constantin, N.K. Schuurman, J.K. Vermunt, A general Monte Carlo method for sample size analysis in the context of network models, Psychol Methods (2023 [Online ahead of print], 10.1037/met0000555).

[42] F.D. Schönbrodt, M. Perugini, At what sample size do correlations stabilize?, Journal of Research in Personality 47(5) (2013) 609–612.

[43] M.W. Petersen, A. Schroder, T. Jorgensen, E. Ornbol, T.M. Dantoft, M. Eliasen, B.H. Thuesen, P. Fink, The unifying diagnostic construct of bodily distress syndrome (BDS) was confirmed in the general population, J Psychosom Res 128 (2020) 109868.

[44] Y.M.J. Goertz, A.M.J. Braamse, M.A. Spruit, D.J.A. Janssen, Z. Ebadi, M. Van Herck, C. Burtin, J.B. Peters, M.A.G. Sprangers, F. Lamers, J.W.R. Twisk, M.S.Y. Thong, J.H. Vercoulen, S.E. Geerlings, A.W. Vaes, R. Beijers, M. van Beers, A. Schols, J.G.M. Rosmalen, H. Knoop, Fatigue in patients with chronic disease: results from the population-based Lifelines Cohort Study, Sci Rep 11(1) (2021) 20977.

[45] A. Hinz, J. Ernst, H. Glaesmer, E. Brähler, F.G. Rauscher, K. Petrowski, R.D. Kocalevent, Frequency of somatic symptoms in the general population: Normative values for the Patient Health Questionnaire-15 (PHQ-15), J Psychosom Res 96 (2017) 27–31.

[46] A. Smakowski, P. Husing, S. Volcker, B. Lowe, J.G.M. Rosmalen, M. Shedden-Mora, A. Toussaint, Psychological risk factors of somatic symptom disorder: A systematic review and meta-analysis of cross-sectional and longitudinal studies, J Psychosom Res 181 (2024) 111608.

[47] W. Rief, G. Rojas, Stability of somatoform symptoms--implications for classification, Psychosom Med 69(9) (2007) 864–9.

[48] F.H. Creed, I. Davies, J. Jackson, A. Littlewood, C. Chew-Graham, B. Tomenson, G. Macfarlane, A. Barsky, W. Katon, J. McBeth, The epidemiology of multiple somatic symptoms, J Psychosom Res 72(4) (2012) 311–7.

[49] T. Chalder, C. Willis, “Lumping” and “splitting” medically unexplained symptoms: is there a role for a transdiagnostic approach?, J Ment Health 26(3) (2017) 187–191.

[50] M. Lehmann, N.J. Pohontsch, T. Zimmermann, M. Scherer, B. Löwe, Diagnostic and treatment barriers to persistent somatic symptoms in primary care - representative survey with physicians, BMC Fam Pract 22(1) (2021) 60.

[51] A.H.J. Sahm, M. Witthoft, J. Bailer, D. Mier, Putting the Vicious Cycle to the Test: Evidence for the Cognitive Behavioral Model of Persistent Somatic Symptoms From an Online Study, Psychosom Med 86(6) (2024) 569–575.

