## Supplementary Figure S1 for "Mapping the Network of Persistent Somatic Symptoms Across Diseases: A Longitudinal Analysis from the SOMACROSS Research Unit"

**Supplementary Figure S1: Stability and accuracy analysis of the baseline symptom network**

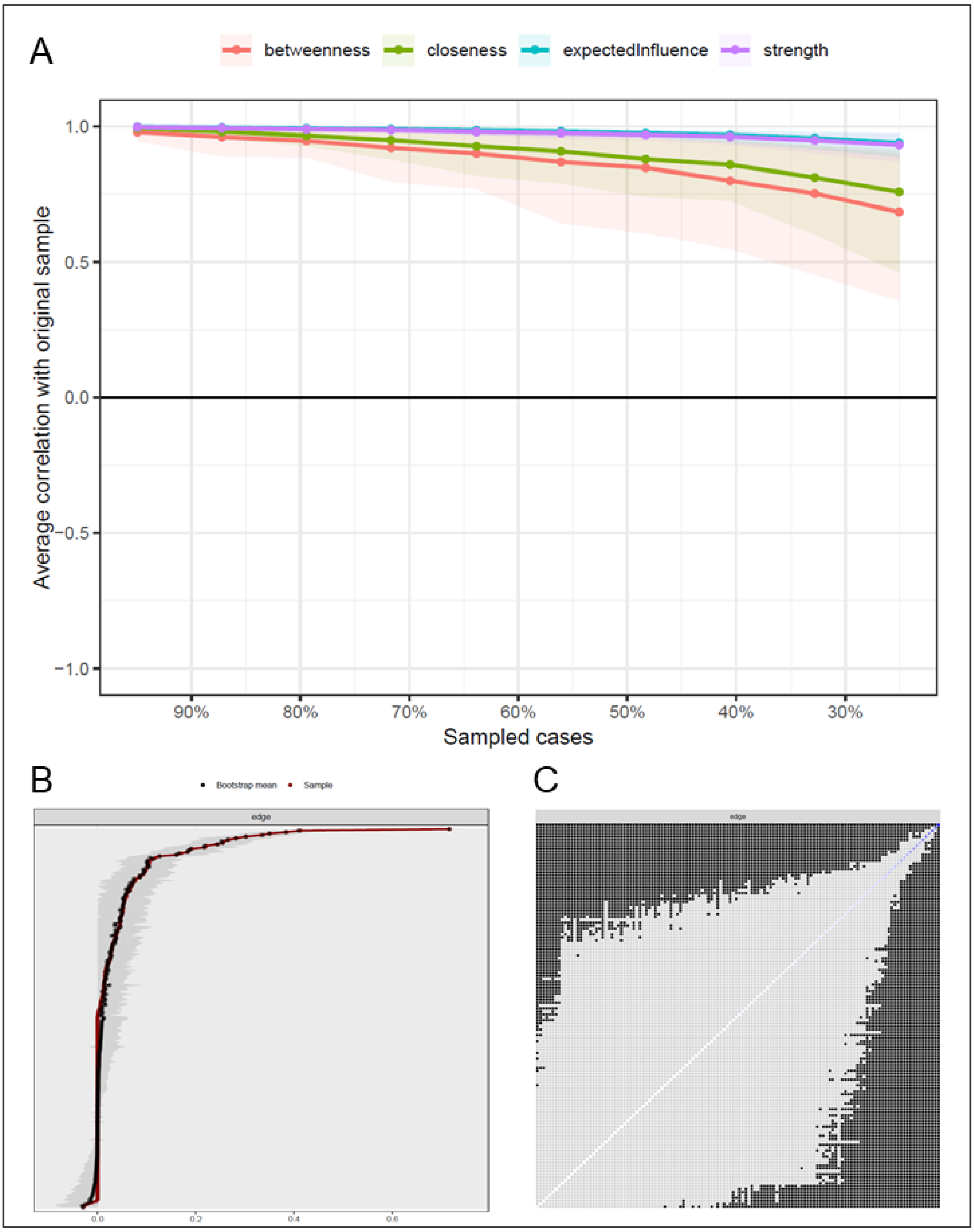


Stability and accuracy of the baseline symptom network. Data include z-standardized scores on the PHQ-15, modified Pain Disability Index, and EURONET-SOMA symptom items from N = 1061 patients (baseline assessment). All panels refer to the network estimated from these baseline data. **(A)** Correlation stability (CS) analysis for four node centrality metrics (strength, expected influence, closeness, and betweenness) obtained via a case-dropping subset bootstrap. In this plot, each colored curve shows the mean correlation between a centrality metric computed on subsampled data and the same metric computed on the full data set; shaded bands denote the 95% confidence interval around each curve. The CS coefficients (correlation stability coefficients) for strength and expected influence reach 0.75, indicating good stability of these two metrics under sample reduction. **(B)** Accuracy of the estimated edge weights assessed by non-parametric bootstrapping. For each network edge, the distribution of bootstrap estimates (based on 1000 iterations) is plotted: the red dot marks the edge-weight estimate from the original data set, and the black dot indicates the mean of the bootstrap distribution. The horizontal spread of the black dots reflects the variability (precision) of each edge-weight estimate – narrower spreads indicate more precise (i.e. accurate) estimation. (**C**) Results of the bootstrapped difference tests for edge weights. Black squares mark pairs of edges that differ significantly ($\alpha = 0.05$) according to the bootstrap difference test (i.e. their confidence interval for the difference does not include zero); the absence of a square indicates no significant difference between those edges. All analyses were conducted in R using the *bootnet* and *qgraph* packages, with 1000 bootstrap iterations for both stability and accuracy assessments.
