## Supplementary Figure S2 for "Mapping the Network of Persistent Somatic Symptoms Across Diseases: A Longitudinal Analysis from the SOMACROSS Research Unit"

**Supplementary Figure S2: Stability and accuracy analysis of the symptom network at 6-month follow-up**

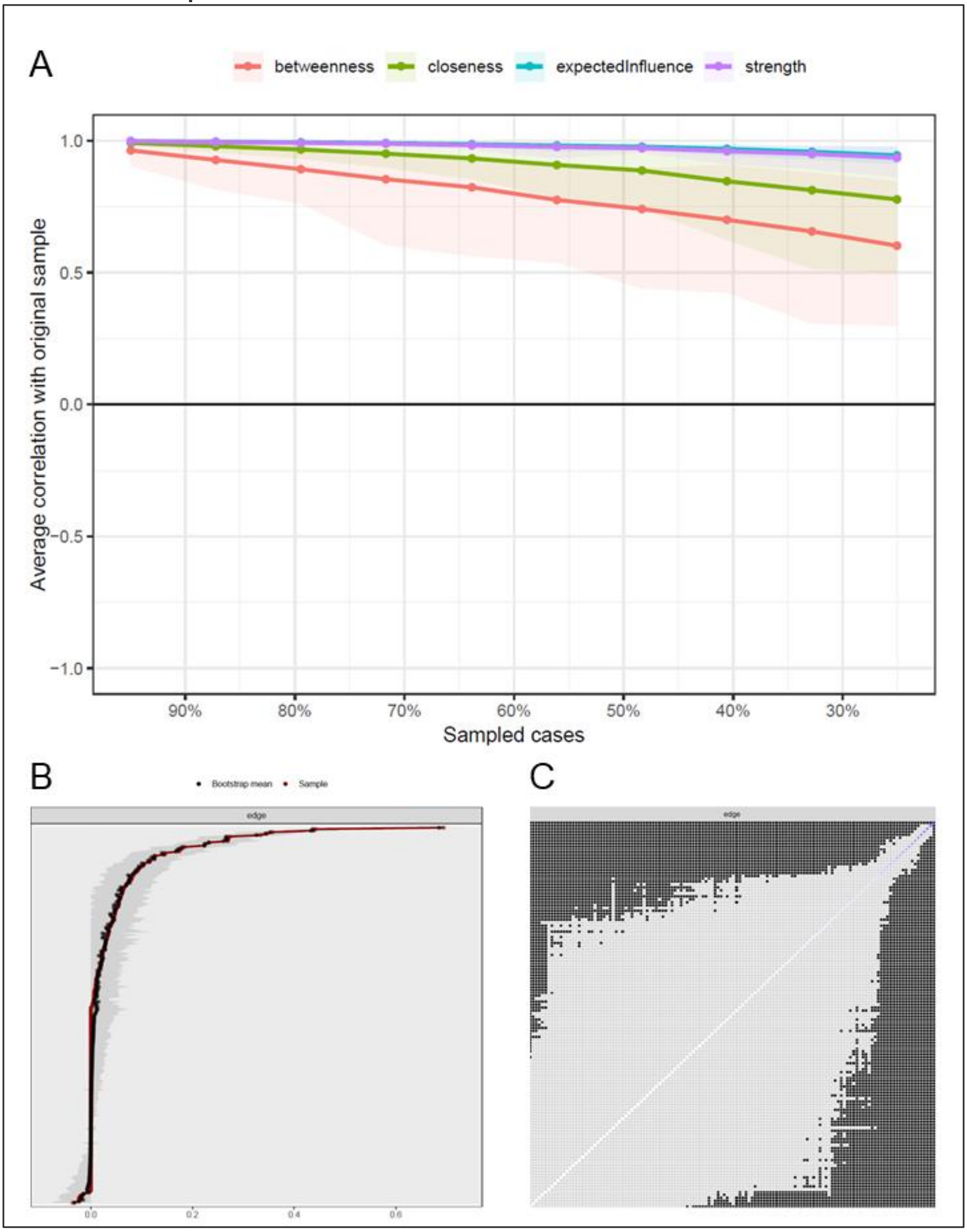


Stability and accuracy of the symptom network (six-month follow-up model) based on z-standardized PHQ-15, modified Pain Disability Index, and EURONET-SOMA items (N = 1134). **(A)** Correlation stability (CS) analysis for four node centrality metrics (strength, expected influence, closeness, and betweenness) obtained via a case-dropping subset bootstrap. In this plot, each colored curve shows the mean correlation between a centrality metric computed on subsampled data and the same metric computed on the full data set; shaded bands denote the 95% confidence interval around each curve. The CS-coefficients for strength and expected influence reach 0.75, indicating that these centrality indices remain highly stable even when a large proportion of cases is dropped. **(B)** Accuracy of the estimated edge weights assessed by non-parametric bootstrapping. For each network edge, the distribution of bootstrap estimates (based on 1000 iterations) is plotted: the red dot marks the edge-weight estimate from the original data set, and the black dot indicates the mean of the bootstrap distribution. The horizontal spread of the black dots reflects the variability (precision) of each edge-weight estimate – narrower spreads indicate more precise (i.e. accurate) estimation. (**C)** Results of the bootstrapped difference tests for edge weights. Black squares mark pairs of edges that differ significantly ($\alpha = 0.05$) according to the bootstrap difference test (i.e. their confidence interval for the difference does not include zero); the absence of a square indicates no significant difference between those edges. All analyses were conducted in R using the *bootnet* and *qgraph* packages, with 1000 bootstrap iterations for both stability and accuracy assessments.
